# Measles RNA detection in wastewater solids

**DOI:** 10.1101/2025.07.18.25331801

**Authors:** Abigail P. Paulos, Alessandro Zulli, Bridgette Shelden, Dorothea Duong, Alexandria B. Boehm, Marlene K. Wolfe

**Author notes:** Authors to whom correspondence should be addressed: Marlene Wolfe, Alexandria Boehm.

## Abstract

Measles incidence has increased in recent years as vaccination rates have dropped globally. However, there are challenges in surveillance of measles; measles presents similarly to other diseases and can be misdiagnosed. The lag between infectivity and symptom onset also poses a challenge for surveillance, as measles is highly infectious and significant transmission can occur before case identification. Wastewater monitoring of measles RNA could help to fill gaps in clinical surveillance. In this study, we developed a novel assay to detect wild-type measles virus in wastewater; through both *in* silico and *in* vivo tests, we demonstrated assay specificity and sensitivity. We conducted both retrospective and prospective monitoring in a sewershed adjacent to ongoing outbreak areas in the region in the United States from December 2024 – May 2025. In total, 11 of 105 (10.5%) of samples were positive for measles with a median concentration of 6,900 gene copies per dry gram of wastewater solids. Overall, we demonstrate that measles is detectable in wastewater during an ongoing outbreak and that wastewater monitoring of measles can result in early warning over clinical surveillance.

## Introduction

Measles is a highly infectious but vaccine-preventable disease, yet global incidence has increased as vaccine coverage has declined since the COVID-19 pandemic.^1^ The measles virus (recently renamed to *Morbillivirus hominis*) is a single-stranded, negative sense RNA paramyxovirus of the genus *Morbillivirus.*^2^ The most common symptoms include fever, rash, cough, rhinitis, and conjunctivitis.^2^ Measles has an average basic reproductive number (R_0_) between 12 – 18, making it one of the most infectious viruses identified.^3^ A vaccine for measles was first licensed in 1963; following the introduction of the vaccine, measles incidence in the

U.S. decreased by more than 95%.^2^ The vaccine is highly effective; ∼95% of those who receive one dose and >99% of those who receive two doses show evidence of measles immunity.^2^ However, because measles is highly infectious, vaccine coverage (2 full doses) of >=95% is needed to prevent outbreaks.^2^ Vaccine coverage, both in the U.S. and globally, has fallen below this threshold in many locations since the COVID-19 pandemic.^1,4,5^

Falling vaccination rates have resulted in an increase in measles cases worldwide since the COVID-19 pandemic, with large outbreaks reported in 37 countries in 2022.^1^ In the U.S., where the national vaccination rate is 90.8%,^6^ over 1,300 cases have been reported in 2025 across 40 US states and territories as of July 16, 2025, with 29 outbreaks (3+ associated cases) identified.^7^ Ninety-six percent of cases are among unvaccinated or under-vaccinated individuals, and 13% of cases have resulted in hospitalizations while three deaths have been reported.^7^

Clinical surveillance for measles is limited by disease presentation, test availability, and healthcare-seeking behavior. The symptoms of measles are similar to those of other diseases caused by the dengue, Zika, or Parvovirus B19 viruses, and during times of low incidence, measles can be hard to differentiate from other similar diseases.^8^ In low-resource settings, availability of laboratory reagents and trained staff can make clinical testing difficult to regularly achieve. In 2022, only half of 144 WHO countries met the measles surveillance target, indicating that enhanced surveillance is needed.^1^ A further limitation of clinical testing is that only cases that seek healthcare will be tested and reported, and milder cases may not be included in case counts.

Wastewater monitoring for measles RNA has the potential to fill in surveillance gaps and provide critical information on measles virus prevalence globally. Measles RNA is shed in urine, saliva, and sputum;^9^ shedding in feces is currently unknown and requires further investigation, especially given that measles results in systemic infection and diarrhea is a symptom in 1 in 10 measles cases.^2,10^ Measles virus RNA partitions preferentially into solids and can persists for days to weeks in wastewater.^11^ Wild-type measle RNA has been detected in wastewater in Chicago,^11^ Texas,^12,13^ Belgium,^14^ the Netherlands,^15^ and Switzerland,^16^ and the vaccine strain (Genotype A) RNA in Ontario wastewater.^17^ Most detections of measles RNA have been at concentrations near method detection limits, with measles RNA detectable but in low concentrations in wastewater even during outbreaks. A key challenge in wastewater monitoring for measles has been the differentiation of the vaccine strain and wild-type measles RNA; the vaccine contains a live, attenuated virus that can be shed in recently vaccinated individuals and is genetically similar to the wild-type strains.^17^ Because outbreaks often result in increased vaccination efforts, differentiating between vaccine and wild-type measles is critical in understanding outbreak progression.

In this study, we (1) developed a novel assay specific to wild-type measles and (2) evaluated the use of this assay for outbreak monitoring by applying it to wastewater testing in an area adjacent to a significant measles outbreak.

## Methods

### Measles assay design

We developed an assay specific to wild-type (WT) measles by modifying an assay previously published by Roy et al. that targets the 3’ region of the measles N gene and was designed to detect only the vaccine strain of measles^18^. We aligned the primer and probe sequences of the Roy et al. assay to 97 measles sequences downloaded from National Center for Biotechnology Information (NCBI) in March 2025 (43 genotype B3 and 54 genotype D8) and to the Edmonstron-Enders and Edmonston-Zagreb vaccine sequences from NCBI Virus using the cloud-based platform Benchling. Simple majority, 75% majority, and 90% majority consensus sequences for both the B3 and D8 genotype were generated in R. We used in silico modeling with RStudio Bioconductor (V3.21) and Benchling software to identify modifications in the probe and reverse primer sequences that would make the assay specific to circulating WT measles genotypes B3 and D8 rather than the vaccine sequences. Based on this analysis, two changes were made, reflected in Table S1 and further described in the Supplementary Appendix.

The ability of the assay to distinguish between wild type and vaccine strains was further confirmed by downloading available sequences from NCBI for the D3, D4, D5, D7, C2, and B2 genotypes and comparing the forward primer, reverse primer, and probe sequences to those genotype sequences as well. The newly designed assay (hereafter, “modified Roy et al.” assay) uses a forward primer (AGGATGAGGCGGACCARTACTT), reverse primer (CRATATCTGAGATTTCCTTGTTCTC), and probe (CATGATGATCCAAGTAGTAGTGA) (also shown in Table S1). We used NCBI BLAST to confirm specificity of the modified Roy et al. assay to WT genotypes B3 and D8. Specificity was checked through an exclusionary BLAST that excludes the intended assay target, allowing for the determination of any potential off target amplification.

The modified Roy et al. assay was tested in vitro against nucleic acids from a panel of respiratory pathogens including coronaviruses, influenza viruses, and several bacterial pathogens (Figure S1). Nucleic acids were extracted and purified using commercially available kits as described below for the wastewater solids samples and then used neat as template in droplet digital 1-step droplet digital RT-PCR assays. The assay was run in a single well using the cycling conditions and post processing using a QX200 (Biorad) droplet reader in singleplex. No template RT-PCR negative controls were included on each plate. Nucleic acid sequences representing genotypes B3 and D8 (WT, gene blocks purchased from IDT, Coralville, IL, Table S1) and genotype A (vaccine strain, purchased from ATCC, VR-24D) were serially diluted in sterile, nucleic-acid free water to achieve template concentrations between 0 and 200 copies per reaction and run in triplicate using droplet digital RT-PCR using the modified Roy et al. assay.

### Testing of wastewater solids for measles RNA

Wastewater solids from two neighboring wastewater treatment plants (WWTPs) were tested for measles RNA. Three samples per week collected between 12/29/24 and 1/14/25 (month/day/year format) and two samples per week collected between 1/19/25 and 4/22/25 were retrospectively tested for measles RNA at each site. Prospective testing of 3 samples/week was conducted between 4/27/25 and 5/13/25. The two WWTPs (hereafter referred to as “North” and “South”) provided 24-h composited influent from sanitary sewer systems; the North services 60,000 and the South 140,000 residents.

Samples were collected using sterile technique by WWTP staff and shipped overnight at 4°C to the laboratory where processing began within 48 h of sample receipt.

Sample pre-analytical processing and nucleic-acid extraction and purification methods are provided in detail elsewhere.^19^ Samples were allowed to sit for 10-15 min and a serological pipette was used to aspirate the settled solids into a falcon tube. Settled solids were centrifuged at 24,000_□_×□g for 30□min at 4□°C to dewater the solids. The supernatant was then aspirated using a vacuum and discarded. A 0.5-to 1-g aliquot of the dewatered, wet solids was dried at 110□°C for 19 to 24□h to determine its dry weight.

We extracted and purified nucleic acids from the solids using Chemagic Viral DNA/RNA 300 Kit H96 (PerkinElmer, Shelton, CT) followed by inhibition removal (Zymo OneStep PCR Inhibitor Removal Kit, Irvine, CA); precise methods are provided on protocols.io^20^ and in other publications.^19^

For samples tested prospectively, the nucleic acids were immediately used neat as template in RT-droplet digital PCR (RT-ddPCR) format and tested for measles RNA using the modified Roy et al. assay . For samples tested retrospectively, nucleic acids were stored at -80°C for 1-3 months before being tested for measles RNA using the assay. All samples tested retrospectively were also tested for SARS-CoV-2 RNA, and the SARS-CoV-2 RNA concentrations measured in the samples after months of storage were compared to those measured immediately without any storage to assess potential for RNA degradation during storage (see SI for details). Bovine coronavirus (BCoV) vaccine was spiked into all samples and measured to assess RNA recovery during the pre-analytical and nucleic-acid extraction and purification processes. See the Supporting Information (SI) for further details on RT-ddPCR reaction chemistry and cycling conditions and processing of machine output.

For each site, we used a Mann-Kendall trend test as a non-parametric approach to assess whether there was any upward or downward monotonic trend in concentrations

## Results

### Assay sensitivity and specificity and QA/QC

*In silico* analyses indicated that the modified Roy et al. assay) is specific to all known and circulating WT measles variants and is not expected to amplify nucleic acids from the vaccine. Notably, since 2018, only B3, D4, D8, and H1 genotypes have been detected in infected individuals globally,^21^ of which only H1 is unlikely to be distinguished by this probe because of additional mismatches on the probe. *In vitro* specificity testing showed no cross reactivity with other respiratory viruses and select bacterial pathogens (Figure S2). *In vitro* sensitivity testing of the modified Roy et al. assay detected both B3 and D8 but did not produce any positive droplets for the vaccine nucleic-acids. The modified Roy et al. assay detected the intended targets (B3 and D8) across all tested concentrations.

The lowest concentration at which all 3 replicates were positive was 6.25 gene copies per reaction for D8, and 25 gene copies per reaction for B3 (Figure S3). The modified Roy et al. assay produced zero positive droplets for all concentrations of the vaccine strain control. Results support the specificity of the assay to genotypes B3 and D8.

Across all experiments, positive and negative controls performed as expected; no amplification was detected in negative controls and all positive controls were positive. Recovery of spiked BCoV RNA across the samples was 1.18 (median, interquartile range (IQR) = 0.64 and 1.72); recovery higher than 1 is due to spiked BCoV RNA measurement uncertainty. SARS-CoV 2 RNA concentrations measured in the samples used for the retrospective testing (and stored for 1-3 months and subjected to a single freeze thaw) were compared to SARS-CoV-2 RNA concentrations in the same samples as measured immediately, without storage. The ratio of concentrations measured in stored versus not stored samples was 1.54 (median, IQR = 1.24 - 1.91) suggesting limited degradation of RNA during the storage time.

### Wastewater results

Measles RNA was detected in 11/105 samples (10.5%) tested; 5 from the North site and 6 from the South site. Of the samples that tested positive, concentrations ranged from 3,929 cp/g to 52,445 cp/g (median 6,900 cp/g). The highest concentration observed was in the first sample that tested positive in Jan 2025 at the North site. There was no significant trend in concentrations of measles RNA in wastewater over time at either site (North p = 0.44, South p = 0.67). Data are available at https://purl.stanford.edu/jm571qx0675.

## Discussion

This study describes the design and application of a wild-type specific assay for the detection of measles viral RNA in wastewater, and demonstrates that this approach can be used to detect measles in wastewater during an active outbreak in the region. We found that an assay specific to wild-type measles (and thus not cross-reacting with any RNA from vaccine strains) was able to detect measles in wastewater from two treatment plants in an area with several reported cases during the study period and adjacent to larger outbreaks.

Measles RNA was first detected in wastewater on 1/7/25, just over a week before the first case in the United States in 2025 was confirmed during the week of 1/20/2025 in Texas. Detections throughout the study period were sporadic, without a pattern of increasing or decreasing concentrations over time. While there were cases reported during April and May 2025 of the study period, there was no sustained outbreak reported in this specific community. Overall, these results suggest that wastewater testing may be able to provide an early indicator of measles circulating in the region and is sensitive to low levels of cases. However, we were not able to observe the association with cases over a sustained outbreak period in the area associated with a treatment plant. Because there were few cases in the area of the treatment plants during the study, and it is suspected that reports may not capture all the cases that occurred during the study time period in the United States, it is difficult to quantify the relationship between wastewater concentrations of measles RNA and the number of cases in the community.

This study demonstrates that measles wild type RNA is detectable in wastewater during a regional outbreak of measles, and that the detections occurred during an active outbreak and prior to the identification of cases in the immediate area.

## Data Availability

Wastewater data are available at https://purl.stanford.edu/jm571qx0675.

## Acknowledgements

We thank the participating wastewater treatment plants for their samples for the project. This work was supported by a gift from the Sergey Brin Family Foundation to ABB.

---

**Fig 1.**
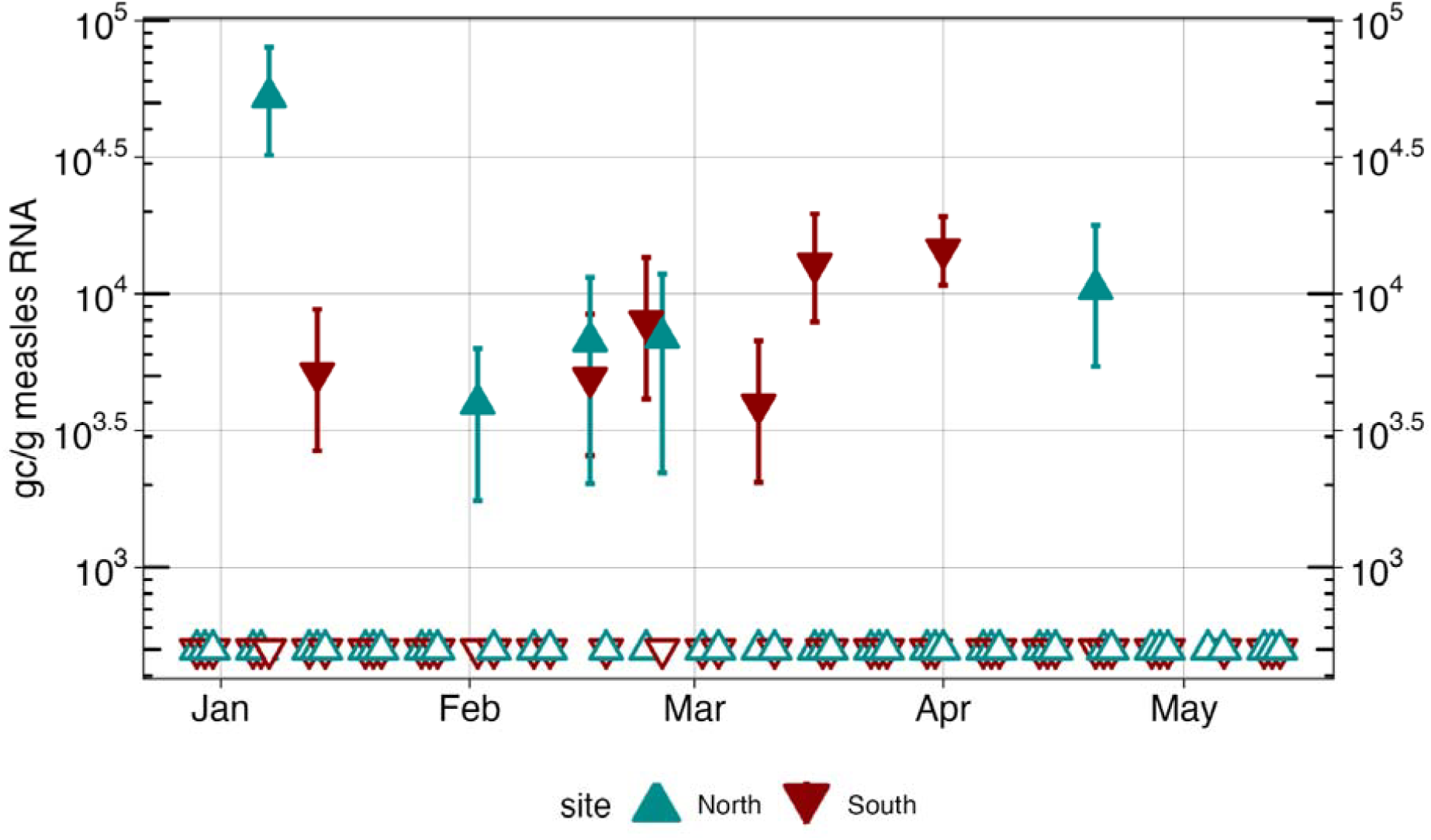
Wastewater measurements of measles RNA. Measles RNA measured in wastewater in cp/g wastewater solids at two WWTPs. Concentrations measured are indicated by filled triangles with the 68% confidence interval represented by bars. Samples with no detection are indicated by open triangles.

