## Supplementary Information for "Measles RNA detection in wastewater solids"

**Measles detection in wastewater solids during an outbreak**

**Additional details on assay design.**

The forward primer from the Roy et al assay was changed to have a degenerate base (R) to allow for simultaneous amplification of both the B3 and D8 genotypes. The probe was changed at position 14, replacing a T with a G. This single nucleotide distinguishes the vaccine strain from the B3, D4, and D8 genotypes circulating worldwide.^20^ Note that although the original Roy et al. assay used a locked nucleic acid probe, the modified assay Roy et al assay does not.

##### Table S1. Modified Roy et al. assay.

| Probe | CATGATGATCCAA**G**TAGTAGTGA |
| --- | --- |
| Forward Primer | AGGATGAGGCGGACCA**R**TACTT |
| Reverse Primer | CRATATCTGAGATTTCCTTGTTCTC |

Bold and underline indicates change from Roy et al. assay

**Additional details on molecular analysis of wastewater samples.** Briefly, solids are isolated from wastewater samples, as described in the main text, and re-suspended in a buffer at a low enough concentration so as to minimize inhibition (75 mg/ml) and homogenized.^1^ Six aliquots of supernatant from each homogenized sample were then subjected to nucleic-acid extraction and purification (Chemagic Viral DNA/RNA 300 Kit H96, PerkinElmer, Shelton, CT), and inhibitor removal (Zymo OneStep PCR Inhibitor Removal Kit, Irvine, CA). Dry weight was determined using another aliquot of solids and drying in an oven.^1^

For the prospective testing, nucleic-acids from the 6 aliquots of each sample were each used used neat as template in 10 replicate droplet digital 1-step RT-PCR (RT-ddPCR) reaction wells to measure measles RNA using the modified Roy et al. assay (Table 1). Nucleic acids from 4 of the aliquots were run in duplicate to obtain the 10 replicate wells. The measles assay was run in multiplex with assays for Dengue virus RNA types 1, 2, 3 and 4 the results of which are not reported herein. The modified Roy et al. assay was run using a probe labeled with the fluorescent molecule fluorescein amidite (FAM).

For retrospective testing, the nucleic-acids from the 6 aliquots of each sample were used as template in 6 replicate 1-step RT-ddPCR reaction wells to measure measles RNA using the modified Roy et al. assay (Table 1). The measles assay (probe labeled using FAM) was run in duplex with an assay for SARS-CoV-2 N gene (probe labeled with hexachlorofluorescein, HEX).^1^

For both retrospective and prospective testing, negative and positive RT-PCR controls were included on each plate (n=2 and 1 wells, respectively). Synthetic gene blocks (IDT, Coralville, IA) controls were used as positive controls for B3 and D8 (Table S1) and negative controls used molecular grade water.

Negative extraction controls and viral RNA recovery controls were also run on the samples. The buffer used for suspending the wastewater solids prior to nucleic-acid extraction was spiked with bovine coronavirus (BCoV) vaccine during the sample processing steps, and nucleic-acid extracts from each sample were assayed for BCoV RNA following methods described elsewhere to calculate BCoV RNA recovery (measured BCoV divided by added BCoV).^1^ Nucleic-acid extracts from the buffer were also used as template for negative extraction controls as they are not expected to contain measles or SARS-CoV-2 RNA targets. The negative and positive controls were run in separate plates that assayed a wide variety of pathogen biomarkers measured prospectively (within 48 h of sample receipt in the lab), as described by Boehm et al.^1^

Digital droplet RT-PCR was performed on 20 μl samples from a 22 μl reaction volume, prepared using 5.5 μl template mixed with 5.5 μl of One-Step RT-ddPCR Advanced kit for Probes (catalog no. 1863021; Bio-Rad), 2.2 μl reverse transcriptase, 1.1 μl dithiothreitol (DTT), and primers and probes at a final concentration of 900 nM and 250 nM, respectively. Droplets were generated using the AutoDG Automated Droplet Generator (Bio-Rad).

PCR was performed using Mastercycler Pro (Eppendorf, Enfield, CT) thermocycler with the following protocol: reverse transcription at 50 °C for 60 min, enzyme activation at 95°C for 5 min, 40 cycles with 1 cycle consisting of denaturation at 95°C for 30 s and annealing and extension at 59°C for 30 s, enzyme deactivation at 98 °C for 10 min, and then an indefinite hold at 4°C. The ramp rate for temperature changes was set at 2°C/s, and the final hold at 4°C was performed for a minimum of 30 min to allow the droplets to stabilize. Droplets were analyzed using the QX200 (retrospective analyses) or QX600 (prospective analyses) Droplet Reader (Bio-Rad). All liquid transfers were performed using the Agilent Bravo (Agilent Technologies).

A well had to have over 10,000 droplets for inclusion in the analysis. Results from replicate wells were merged for analysis. Concentrations of the targets in wastewater samples are presented as copies per gram dry weight. For a sample to be scored as a positive, there had to be at least 3 positive droplets across the merged wells. The lowest measurable concentration is approximately 1000 copies/g dry weight (corresponds to three positive droplets across 6 to 10 merged wells). Errors are reported as standard deviations of the measurements as obtained from vendor’s software.

**Additional details related to EMMI guidelines.** The retrospective samples had an average (standard deviation) droplets across the 6 replicate wells of 96,265 (20,913). The volume per droplet, as reported by the manufacturer, is 0.00085 μL. The average (standard deviation) copies per droplet of MeV was 4.5 x 10^-5^ (2.6x10^-4^). For the prospective sampling, the average (standard deviation) droplets was 188,187 (17,139). The average (standard deviation) copies per droplet was 1.1 x 10^-6^ (7.3 x 10^-6^).

For an example fluorescence plot, see Figure S2.

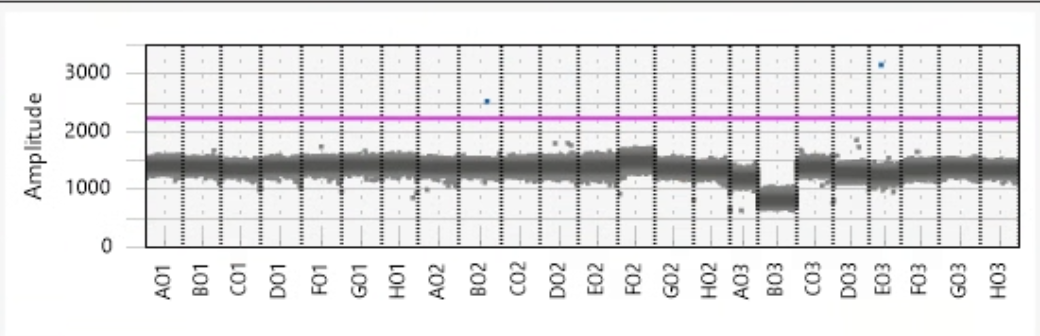

##### Figure S1. Amplitude plot for in vitro specificity testing. The amplitude of fluorescence for each sample is shown in the y-axis. The pink line separates positive (above line) droplets in blue and negative (below line) droplets in grey. From left to right, A01 is Parainfluenza 1, B01 is parainfluenza 2, C01 is parainfluenza 3, D01 is parainfluenza 4, E01 is influenza A H1N1pdm, F01 is influenza A H1, G01 is influenza A H3, H01 is influenza B, A02 is adenovirus 1, B02 is adenovirus 3, C02 is adenovirus 31, D02 is Rhinovirus Type 1A, E02 is RSV A, F02 is SARS-CoV-2, G02 is *M. pneumoniae,* H02 is *C. pneumoniae*, A03 is metapneumovirus 8, B03 is coronavirus HKU-1, C03 is coronavirus 229E, D03 is coronavirus NL63, E03 is coronavirus OC43, F03 is *B. parapertussis*, G03 is *B. pertussis*, H03 is a no template control. One positive droplet is observed in locations B02 and E03, containing adenovirus 3 and coronavirus OC43, but three droplets must be positive to score a sample as positive, so these are negatives. All targets are part of a panel purchased from Zeptometrix (Buffalo, NY) and are inactivated pathogens.

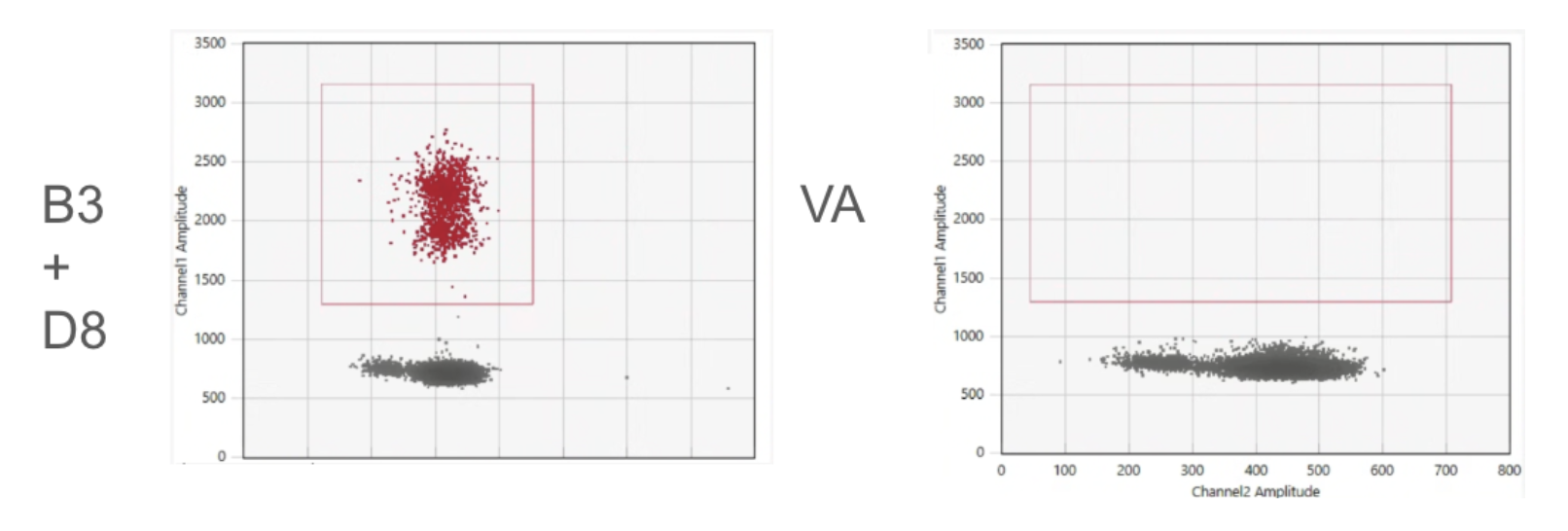

##### Figure S2. Amplitude plot showing droplets from digital PCR testing for sensitivity for the modified Roy et al. assay. Positive droplets are shown in red and negative are shown as grey. The left panel shows results for equal molar concentrations of gblocks for B3 and D8 (approximately 1000 copies of each per reaction) as template. The right panel shows results for vaccines nucleic acids (approximately 1000 copies per reaction). The red boxes indicate the region of the plots where positive droplets are expected based on thresholding.

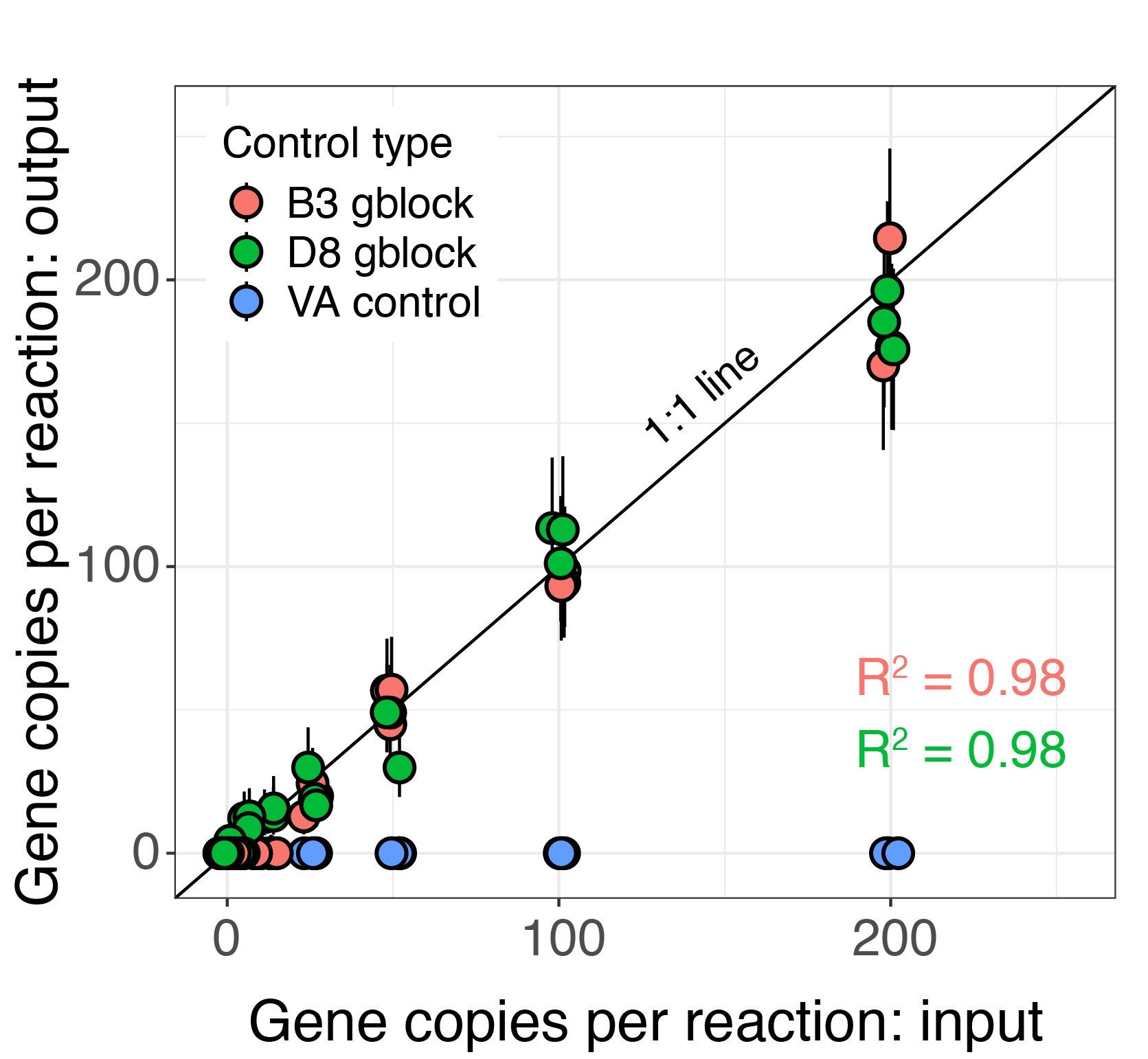

##### Figure S3. Sensitivity testing using the modified Roy et al. assay. Expected concentrations based on preparation of gblock control material and measured concentrations with and without the RT step for digital PCR. Error bars represent the standard deviation. If error bars cannot be seen, they are smaller than the symbols. VA represents the vaccine sequence.

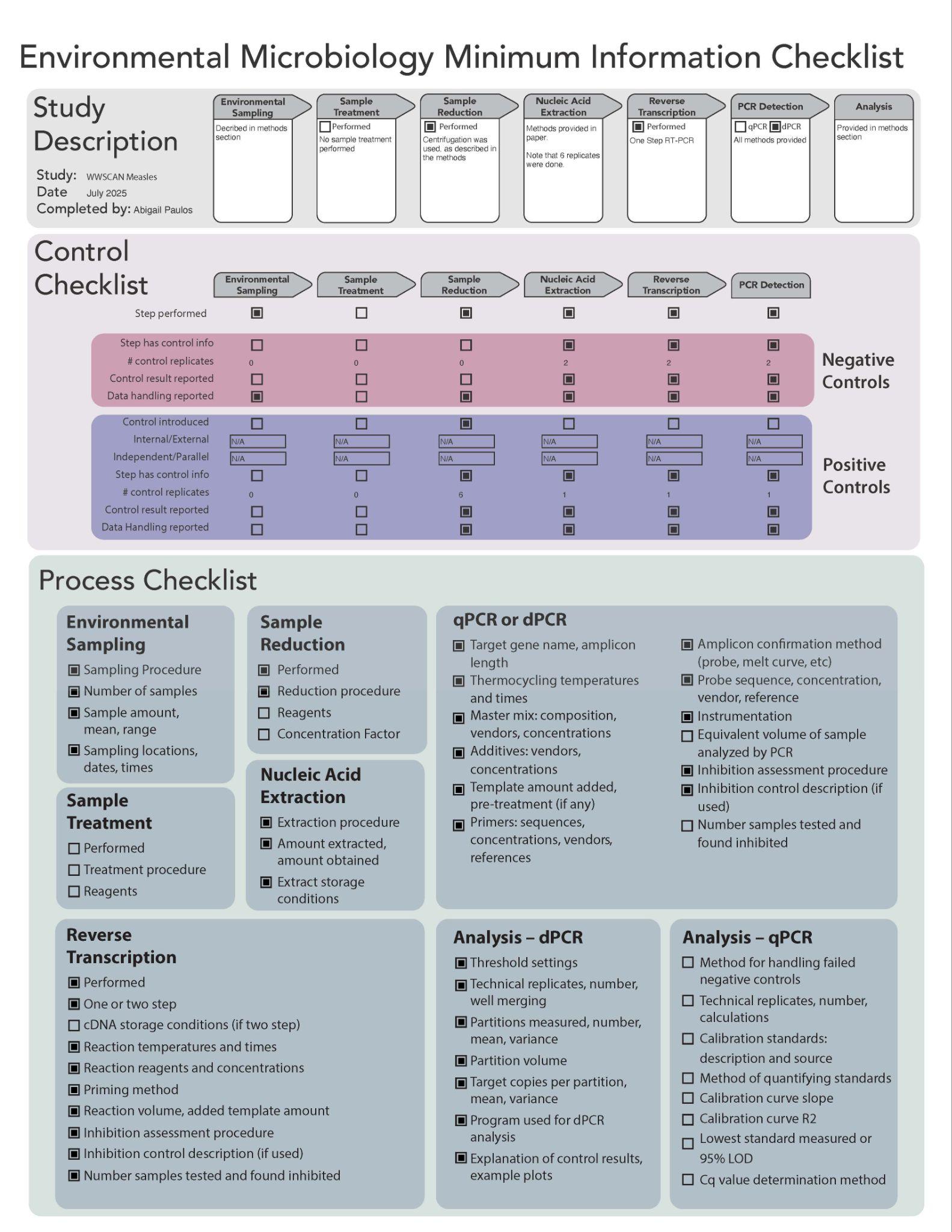

##### Table S1. Gene block sequences for wild type measles genotypes D8 and B3.

| D8 gene block | ATCTGGCCTCACCTTCGCATCAAGAGGTACCAACATGGAGGATGAGGCGGACCAATACTTTTCACATGATGATCCAAGTAGTAGTGATCAATCCAGGTTCGGATGGTTCGAGAACAAGGAAATCTCAGATATCGAAGTGCAAGACCCTGAGGGATTTAACATGATTCTGGGTACCATTCTAGCCCAAATTTGGGTCTTGCTCGCAAAGGCGGTTACGGCCCCAGACACGGCAGCTGATTCGGAGCTAAGAAGGTGGATAAAGTACACCCAACAAAGAAGGGTAGTTGGTGAATTTAGATTGGAGAGAAAATG |
| --- | --- |
| B3 gene block | GCCTTACCTTCGCATCGAGAGGTACTAATATGGAGGATGAGGCGGACCAGTACTTTTCACATGATGATCCAAGTAGTAGTGATCAATCCAGGTTCGGGTGGTTTGAGAACAAGGAAATCTCAGATATTGAAGTGCAAGACCCTGAGGGCTTCAACATGATTCTGGGTACCATCTTAGCTCAAATTTGGGTCTTGCTCGCAAAGGCGGTTACGGCTCCAGACACAGCAGCTGATTCAGAGCTAAGAAGGTGGATCAAATACACCCAACAAAGAAGAGTAGTTGGTGAATT |
